# Infections after routine syndesmotic screw removal – a retrospective quality study

**DOI:** 10.1101/2020.10.30.20222216

**Authors:** Thomas Aleksander Øverby, Rune B. Jakobsen

## Abstract

**Background:** Infections after routine removal of syndesmotic screws are an unwanted complication.

**Methods:** We included all patients treated with syndesmotic screw with routine removal in a five-year period and extracted information regarding fracture characteristics and patient characteristics. Infections were classified as superficial or deep incisional.

**Results:** 1246 patients with ankle fractures were treated surgically in the study period. 343 had routine removals of the syndesmotic screw. We identified 6 cases with infection (1.7%) developing after routine syndesmotic screw removal.

**Conclusions:** We found a low infection rate but when infections did occur, they frequently required surgical revision(s) and long-time antibiotic treatment.

## 1. INTRODUCTION

Ankle fractures are common with an annual incidence of up to 187 per 100.000 persons.^1-3^ Depending on health systems and trauma panorama somewhere between 14 and 72 % of patients with an ankle fracture receive operative treatment.^4^ About 1 in 5 patients with an ankle fracture needing internal fixation will have a concomitant injury to the distal tibiofibular syndesmosis.^5^ The gold standard for treating syndesmotic injury is transsyndesmotic fixation with either one or two metal screws, however no real consensus exists on size of screws, number of cortices or when to start full or partial weight bearing nor is there agreement on if and when screw(s) should be removed.^6, 7, 8^ Suture-buttons are an alternative to screw fixation and have been shown to provide equal or better results but are expensive and in 5-10 % of cases also require removal.^9-12^ The routine in our department is one 4.5 mm quadricortal syndesmotic screw with partially weight bearing allowed after 6 weeks and routine removal of the screw after 12 weeks at the orthopaedic outpatient clinic. Standard radiographic evaluations are performed after 6 weeks and before screw removal at 12 weeks.

Recent publications have reported infection rates of 5 to 9 % resulting from routine removal of syndesmotic screws and suggested that a single dose antibiotic prophylaxis should be given intravenously.^13, 14^ The aim of the present study was to determine the rate of infections after routine removal of syndesmotic screw in our institution to determine whether a change in protocol to routine antibiotic prophylaxis is warranted.

## 2. METHODS

### 2.1. Patients

We included all patients who were surgically treated with syndesmotic screw fixation with subsequent removal at Akershus University hospital in a five-year period (Figure 1). Patients were retrospectively identified by a systematic search in the electronic patient record system for diagnostic codes relating to ankle fractures with concurrent surgical codes relating to ankle surgery and removal of metal. The resulting list was then used to manually examine the patient record to confirm that the patient could be included.

**Figure 1:**
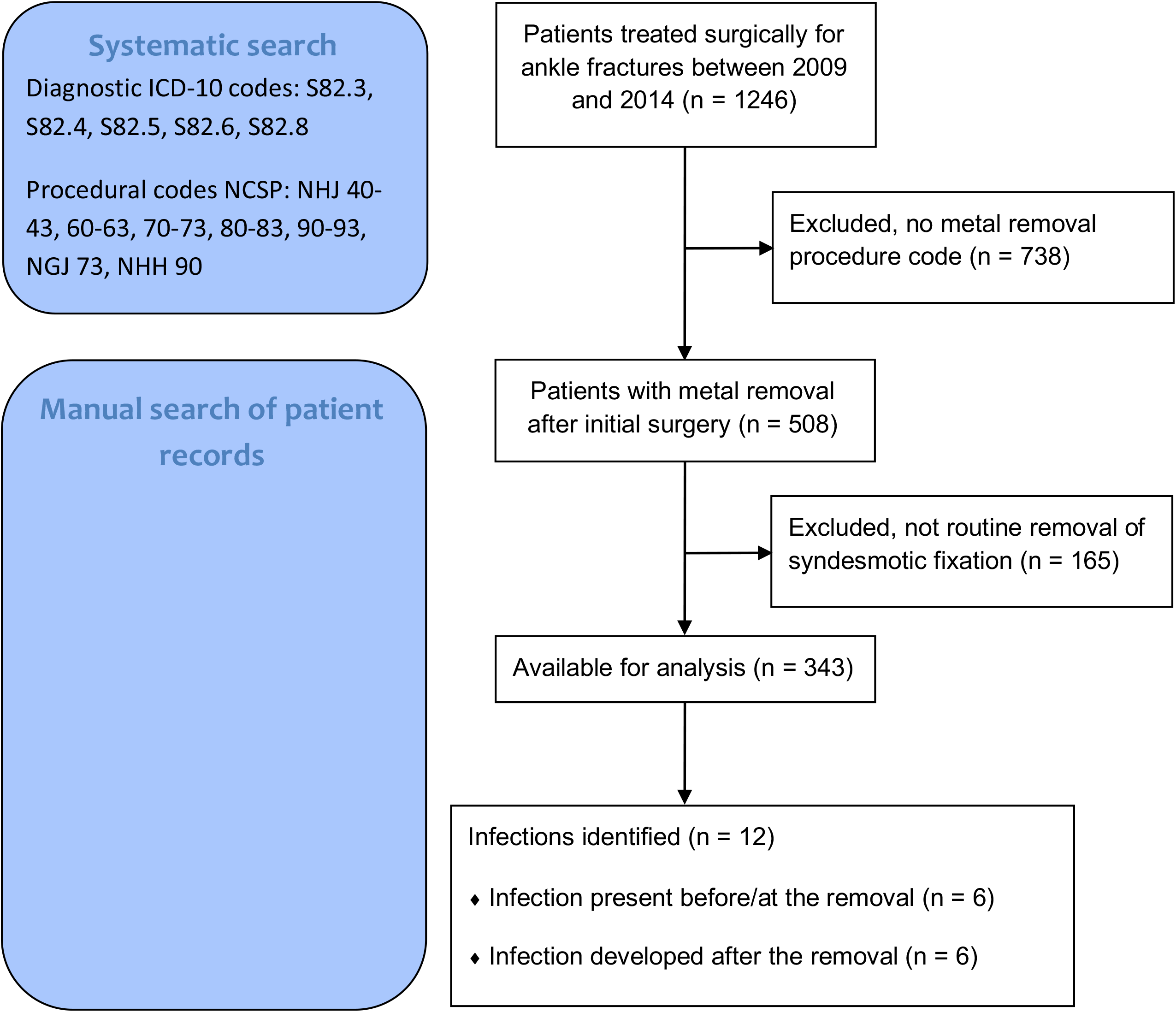
Flow chart of inclusion and identified infections. Flow chart describing the systematic and manual search of patient records with inclusion and exclusion criteria. NCSP: The NOMESCO Classification of Surgical Procedures, ICD-10: International Classification of Diseases 10.

### 2.2. Data extraction

For each patient we extracted the following information from the record and the associated radiographs: fracture characteristics (AO-classification), age, sex, American Society of Anaesthesiologists Physical Status Classification (ASA), body mass index (BMI), smoking and diabetic status, localisation for metal removal (outpatient clinic, operating theatre), metal remaining in the ankle after screw removal, infection diagnosed before, at or in the 12 weeks after routine metal removal either diagnosed at the hospital or documented in the patient record from contact with general practitioners (GPs), antibiotic use (type, dose and length of treatment) and if treatment required surgical revision and/or hospitalisation. We also recorded the microbes identified. Infections were classified as superficial or deep incisional using the Center for Disease Control (CDC) definition.^15, 16^ We used a web-based questionnaire to collect the data.

### 2.3. Statistics

We used Fischer’s Exact Test to asses for significant differences regarding sex, smoking, diabetes status and remaining metal and Student’s unpaired t-test for comparing means. Statistical significance was assumed for p <0.05. Statistical analysis was performed using SPSS 24.0 (SPSS Inc., Chicago IL). For further details about the data sampling, see Figure 1. The project was approved the by the Board of Research at the Department of Orthopedic Surgery and the Data Protection Official of our institution (#16-085). The study was undertaken as an internal quality assurance project and did not involve recontacting patients. As such it did not fall under The Health Research Act and did not require further approval.

## 3. RESULTS

1246 patients with ankle fractures were treated surgically in the study period. 508 patients subsequently had metal removed of which 343 were routine removals of the syndesmotic position screw. Mean age at time of removal was 47.9 years, 56 % were males, and mean BMI was 28.6. 61 % of patients had remaining metal after screw removal. Detailed patient characteristics are listed in Table 1 and fracture characteristics in Table 2. In 330 patients, the syndesmotic fixation removed was a single 4.5 mm quadricortical screw and in 13 patients two 3.5 mm tricortical screws. None of the patients received prophylactic antibiotics prior to removal. We identified 12 (3.5 %) patients with infections, however 6 (1.7 %) of these were documented in the patient record as present either before or at the time of screw removal, leaving 6 (1.7 %) infections developing after routine removal. All patients with infections after removal had quadricortical fixation with an average time of 90 days between surgery and screw removal which in all cases were performed at the out-patient clinic. Of the infections diagnosed after removal 4 of the infections were superficial and 2 were deep incisional infections. 3 of these patients were hospitalised with intravenously antibiotics and surgical revisions and 3 were treated as outpatient with oral antibiotics only. Details about each of the 6 patients are given in Table 3. One of the patients were initially treated by her GP without consultation with the orthopaedic department, but due to poor response of oral antibiotics, the patient was referred to our institution 77 days after screw removal and treated with intravenous antibiotics, surgical revision and negative pressure wound therapy. This patient was the only one with significant sequela in form of decreased range of motion in the ankle joint. In 5 of the 6 patients *Staphylococcus Aureus* (non-MRSA) was identified as the causing microbe, and in *2 β-haemolytic streptococcus* were identified as well. In the last patient no microbe was identified.

**Table 1.** Patient characteristics.

| Patient characteristics |  |  |  |  |
| --- | --- | --- | --- | --- |
|  | Total<br>(*n*=343) | Without<br>infection<br>(*n*=331) | With infection<br>before/at<br>removal(*n*=6) | With infection<br>after<br>removal(*n*=6) |
| Male | 191 (56%) | 184 (56%) | 2 (33%) | 5 (83%) |
| Age, years | 47.9 (16.8) | 47.6 (16.7) | 63.3 (10.1) | 50.1 (17.5) |
| ASA classification<br>1-2(*n*=302) | 302 (88%) | 292 (88%) | 4 (67%) | 6 (100%) |
| BMI, kg/m<sup>2</sup>(*n*=241) | 28.6 (5.0) | 28.6 (5.0) | 31.5 (7.3) | 28.2 (4.1) |
| Diabetes Mellitus<br>(*n*=340) | 20 (6%) | 19 (8%) | 1 (17%) | 0 (0.0%) |
| Smoking(*n*=340) | 88 (26%) | 83 (36%) | 4 (67%) | 1 (17%) |
| Remaining metal | 210 (61%) | 202 (61%) | 3 (50%) | 5 (83%) |

Values are expressed as mean (SD) or as numbers (%)

**Table 2.** Fracture characteristics.

|  | Total<br>(*n*=343) | Without<br>infection<br>(*n*=331) | With infection<br>before/at<br>removal(*n*=6) | With infection<br>after<br>removal(*n*=6) |
| --- | --- | --- | --- | --- |
| AO type 44-B | 38 (11 %) | 38 (11%) | 0 (0.0%) | 0 (0.0%) |
| AO type 44-C | 133 (39%) | 128 (39%) | 2 (33%) | 3 (50%) |
| Isolated<br>syndesmotic<br>injury | 5 (1%) | 5 (1%) | 0 (0.0%) | 0 (0.0%) |
| Bimalleolar | 106 (31%) | 102 (31%) | 3 (50%) | 1 (17%) |
| Trimalleolar | 61 (18 %) | 58 (18%) | 1 (17%) | 2 (33%) |

Values are expressed as numbers (%)

**Table 3.** Details on the infections identified after routine screw removal.

| Patient | Sex | Agegroup<br>(decade)* | Fracture<br>classification | Time from<br>primary<br>surgery to<br>removal<br>(days) | Time<br>from<br>removal<br>to<br>contact<br>with<br>hospital | BMI | ASA | Smoking<br>(yes/no) | Diabetes<br>(yes/no) | Microbiology | Total<br>number of<br>days<br>treated<br>with<br>antibiotics | Type of<br>antibiotics<br>and number<br>of doses | Comments |
| --- | --- | --- | --- | --- | --- | --- | --- | --- | --- | --- | --- | --- | --- |
| 1 | M | 50's | AO type 44-C | 88 | 11 | 31 | 2 | N | N | - | 10 | 30 doses<br>(dicloxacillin) | Superficial infection. No surgical revision. No bacteria identified. |
| 2 | F | 40's | AO type 44-B<br>(trimalleolar) | 84 | 77 | 25 | 2 | Y | N | *S. Aureus* | 34 | 96 doses<br>(dicloxacillin)<br>40 doses<br>(cloxacillin) | Superficial infection. Treated by GP with oral antibiotics before referral. Surgical revision, metal removal and NPWT. Patient had external fixation before primary surgery, syndesmotic fixation performed 3 days after primary surgery after review of radiographs. |
| 3 | M | 70's | AO type 44-C | 98 | 39 | 33 | 2 | N | N | *S. Aureus, β-haemolytic streptococcus* | 24 | 72 doses<br>(clindamycin) | Superficial infection. No surgical revision. Patient had syndesmotic screw repositioning 4 days after primary surgery after review of radiographs. |
| 4 | M | 30's | AO type 44-C<br>(trimalleolar) | 103 | 10 | 29 | 2 | N | N | *S. Aureus* | 87 | 348 doses<br>(dicloxacillin) | Deep infection. Elevated CRP at admission. Surgical revision, metal removal and NPWT. Patient had external fixation before primary surgery. |
| 5 | M | 60's | AO type 44-B<br>(bimalleolar) | 83 | 6 | 23 | - | N | N | *S. Aureus β-haemolytic streptococcus* | 57 | 228 doses<br>(dicloxacillin) | Deep infection. Elevated CRP and fever at admission. Surgical revisions and NPWT. |
| 6 | M | 20's | AO type 44 C | 89 | 4 | - | 1 | N | N | *S. Aureus* | 45 | 28 doses<br>(cloxacillin)<br>152 doses<br>(dicloxacillin) | Superficial infection. Followed as outpatient. All metal removed when radiographically healed. |

AO: Arbeitsgemeinschaft für Osteosynthesefragen. BMI: Body mass index. ASA: American Society of Anesthesiologists Physical Classification System.
NPWT: Negative pressure wound therapy. \* Age given within decade for privacy

We found no statistically significant differences for the variables in Table 1 between patients with no infection and patients with infection after screw removal.

In total, 994 doses of antibiotics were used to treat the infections in our institution (Table 3). One patient had a short prescription for antibiotics before referral to our hospital, estimated to approximately 50 doses. Mean number of days treating the superficial infections were 28 days, and for the deep infections 72 days. The most frequent antibiotic prescribed was dicloxacillin, but also cloxacillin and clindamycin were used.

## 4. DISCUSSION

Over the 5-year period studied, we found an infection rate of 1.7 % after routine syndesmotic screw removal. Most infections were classified as superficial, yet 3 out of the 6 patients required one or more surgical revisions and all patients were treated with antibiotics for a substantial length of time. Previous retrospective cohorts have found higher wound infection rates of 5 % and 9 % after syndesmotic screw removal.^13, 14^ We have no clear explanation for the lower rate found in our study. The electronic and manual search of the patient record was thorough and extensive. One of the patients with infection had initially been treated at the GP, and it is possible that more patients may have been treated for a superficial infection in the primary health sector without subsequent contact with the orthopaedic department. Arguably, the previous studies were small and only a few infections would amount to a considerable percentage compared to our larger cohort. Our manual search did also allow us to differ between infections developing after the removal and infections already present at the time of removal, which was not commented on in the previous studies.

One previous study has recommended to give prophylactic antibiotics in form of broad-spectrum cefalotin when removing syndesmotic fixation.^13^ This is not unreasonably as it has been shown that an intravenous dose of antibiotics given within 30-60 minutes prior to clean orthopaedic procedures cuts the risk of infection by approximately 50 %.^17-20^ The total doses of antibiotics used to the treat the infections identified in the present study was 994.

Predominantly, this was narrow-spectrum betalactam antibiotics in the form of dicloxacillin and cloxacillin. As methicillin-resistant *Staphyloccus Aureus* (MRSA) is not yet a common pathogen in Norway, dicloxacillin is the recommended therapy for surgical site infections below the knee. This also seems a prudent choice since *Staphyloccus Aureus*, in line with other studies, was found in all patients with infections where a microbe could be identified.^13, 20, 21^ Hypothetizing, the total number of prophylactic doses of the broad-spectrum cefalotin to the patients in this cohort would have amounted to 343, approximately one third of the therapeutic doses of narrow-spectrum antibiotics administered. This would demand significantly more resources in the out-patient clinic establishing intravenous access and stocking necessary antibiotics and other equipment. If this could lead to a decrease of infections by 50% and subsequently a similar decrease of therapeutic antibiotic use this might be justified even with a rate as low as 1.7 %. However, a recent adequately powered randomised placebo-controlled trial found that a single preoperative dose of cefazolin did not reduce the risk of surgical site infection following removal of orthopaedic implants below the knee.^22^ As such a change in protocol at our institution would seem to contain disadvantages by increasing the use of resources generally and specifically of broad-spectrum antibiotics which could drive increased resistance of bacteria to cephalosporins.^23^

One might argue that another strategy could be to selectively identify the patients at risk of developing infection and only administer prophylactic antibiotics to them, however in line with others authors we could not identify significant risk factors associated with developing infection, yet 5 of the 6 patients who developed infection were men consistent with findings in other studies that infection after orthopaedic surgery is more frequent in males.^13, 14, 24^ Limitations of our study is the retrospective design relying fully on what was documented in the electronic patient record which may have lead us to underestimate the infection rate, however in a similar study direct contact with the patients by questionnaire did not reveal additional infections besides the ones already documented in the record.^13^ Another limitation is the lack of a control group, and a more preferable study design would have been a randomised controlled trial. However, a sample size calculation (power = 80 %, alpha = 0.05) with an infection rate of 2 % and an anticipated 50 % decrease by giving prophylactic antibiotics results in sample size of 2318 patient in each group which is not feasible.

## 5. Conclusion

We found a lower infection rate after routine syndesmotic screw removal in a large cohort of patients than found in previous publications. When infections did occur, they frequently required surgical revision(s) and long-time antibiotic treatment. Giving prophylactic antibiotic might reduce the risk of infection but likely not to an extent warranting the increased use of a broader spectrum antibiotic.

## Data Availability

Anonymized dataset is available upon request, please contact the last author.

